# Futility of combining griseofulvin and terbinafine in current epidemic of altered dermatophytosis in India: Results of a randomized pragmatic trial

**DOI:** 10.1101/19007617

**Authors:** Sanjay Singh, Vinayak N Anchan, Radhika Raheja

**Affiliations:** Department of Dermatology and Venereology, Institute of Medical Sciences, Banaras Hindu University, Varanasi

## Abstract

**Background:** Treatment responsiveness of tinea has decreased considerably in recent past in India. We tested effectiveness of oral terbinafine plus griseofulvin versus terbinafine alone in tinea corporis, tinea cruris and tinea faciei in a randomized pragmatic open trial.

**Methods:** One hundred and thirty two microscopy confirmed patients were randomly allocated (ratio 1:1) to two groups, terbinafine (T) and terbinafine plus griseofulvin (T+G). Doses given were as follows: T, oral terbinafine (6 mg/kg/day, maximum 500 mg/day, once daily); T+G, terbinafine (as above) plus oral griseofulvin (children [<18 years] 10 mg/kg/day, adults [18 years or more] 10 mg/kg/day, but not <500 mg and not >1000 mg per day, in two divided doses). Patients were treated for 8 weeks or cure, whichever occurred earlier.

**Results:** At 4 weeks, none of the patients were cured in both groups. At 6 weeks, 1(1.5%) and 4 (6.1%) patients were cured in T and T+G groups, respectively (P=0.417). At 8 weeks, 17 (25.8%) and 19 (28.8%) patients were cured in T and T+G groups, respectively (P=0.845). For cure rate at 8 weeks, number needed to treat (NNT) for T+G (versus T), was 33.

**Conclusions:** Addition of griseofulvin to terbinafine does not increase effectiveness of terbinafine in current epidemic of altered dermatophytosis in India.

## Introduction

Unprecedented changes in the epidemiology, clinical features and treatment responsiveness of dermatophytosis have been noted in recent past in India.^1,2^ Recent data show that oral terbinafine, once a highly effective drug, now has an abysmal cure rate in tinea corporis, tinea cruris and tinea faciei.^3,4^ Decreased effectiveness of terbinafine has been attributed to mutation in the squalene epoxidase gene.^5,6^ Furthermore, there is evidence of decreased effectiveness of other oral antifungal drugs (fluconazole, griseofulvin, itraconazole) also in India.^4^ Importance of finding effective methods of treating tinea cannot be overemphasized.

In the present study, we tested the hypothesis that using a regimen consisting of more than one oral antifungal drugs may produce better treatment outcome. We performed a clinical trial comparing effectiveness of oral terbinafine (T, active control) versus an experimental regimen, terbinafine plus griseofulvin daily (T+G), in an investigator-initiated, two-arm randomized pragmatic open trial.

## Methods

### Setting

The study (registered in Clinical Trials Registry-India, registration number CTRI/2019/06/019778) was conducted at a tertiary care centre after obtaining approval from the Institutional Ethics Committee.

### Sample size

A sample size of 66 patients in each group was determined using expected cure rates of 30% and 60% with the control and experimental regimens, respectively, with type I error rate of 0.05, type II error rate of 0.1 and dropout rate of 20%.^7^

### Selection criteria and enrolment

Patients with tinea corporis, tinea cruris or tinea faciei or a combination of these conditions were included in the trial. Inclusion criteria were (a) clinical suspicion of tinea corporis, tinea cruris or tinea faciei or a combination of these conditions, (b) microscopic confirmation (20% potassium hydroxide [KOH] microscopy), and (c) age 4 to 80 years. Exclusion criteria were (a) presence of any other type(s) of tinea, e.g., onychomycosis, (b) pregnancy, (c) lactation, (d) inability to come for follow-up, (e) history of adverse reaction to terbinafine or griseofulvin, (f) abnormal complete blood count (CBC, for haemoglobin and platelet count the cut-offs were <9 gram/dl and <100,000 per microliter, respectively), liver and renal function tests (LFT, RFT), plasma glucose (fasting and postprandial), glycosylated haemoglobin (HbA1c), fasting lipid profile, urine routine and microscopy, electrocardiogram (ECG), (g) current treatment with drugs likely to cause interaction with terbinafine or griseofulvin, (h) present history of renal, liver or ischemic heart disease, (i) presence of another skin disease at the site of tinea, (j) treatment with terbinafine or griseofulvin in last one month, and (k) patients requiring antihistamines due to other skin diseases. A witnessed, written and informed consent was given by the patients, or by a parent in case of minor patients. All female patients of child bearing age were advised to avoid pregnancy during treatment.

Seven hundred and ten patients with suspected tinea corporis, tinea cruris or tinea faciei or a combination of these conditions attending dermatology outpatient department were assessed for eligibility. A total of 132 patients satisfied the selection criteria and were assigned to one of the two treatment groups by block randomization on the basis of random numbers generated online using Research Randomizer (https://www.randomizer.org/). Each treatment group comprised 66 patients (allocation ratio 1:1). Study was conducted from June 2019 to September 2019. Allocation was concealed using sequentially numbered, sealed, opaque envelopes, which contained the allocation code written on a folded paper, the envelopes were opened after enrolment of the patients. Both random sequence generation and allocation concealment were done by a person unrelated to the trial.

### Intervention

Doses given to the patients were as follows: T (oral terbinafine, 6 mg/kg/day, maximum 500 mg/day, once daily), and T+G (oral terbinafine, 6 mg/kg/day, maximum 500 mg/day, once daily; plus oral griseofulvin (children [<18 years] 10 mg/kg/day, adults [18 years or more] 10 mg/kg/day, but not <500 mg and not >1000 mg per day, in two divided doses)). Patients received the treatment for 8 weeks or cure, whichever occurred earlier. Patients were advised against using any other treatment.

### Follow-up

Following data were recorded for each patient: clinical details including duration of illness, body surface area (BSA) affected, treatment given, results of all investigations as above at baseline, microscopy results, and follow-up data. In the terbinafine group (T), CBC and LFT were done every 2 weeks. In the patients who received terbinafine plus griseofulvin (T+G), following investigations were done every 2 weeks: CBC, LFT; and following every 4 weeks: RFT, and urine routine and microscopy.

The most severe lesion was identified as the index lesion, from which scraping and KOH microscopy was performed at baseline and then at subsequent visits. Patients were followed up at 2 weekly intervals up to a maximum of 8 weeks or cure, whichever occurred earlier.

### Measurement of treatment effect

Cure was defined as occurrence of both clinical cure (complete clearance of lesions) and mycological cure (negative KOH microscopy). Presence of post-inflammatory hyperpigmentation at the site of healed tinea was not considered a feature of tinea. Any of the following was considered treatment failure: (a) no improvement or worsening of the disease in 4 weeks in patient’s assessment, (b) presence of scaling and/or erythema at 8 weeks, and (c) positive microscopy at 8 weeks. Patients who were cured were asked whether they were fully satisfied with the treatment outcome or not. Patients in whom the treatment failed were treated outside the trial with oral itraconazole.

### Measurement of compliance

Patients were asked to bring used strips of tablets and the strips in use at each follow up visit to assess compliance. Compliance was considered to be good if a patient had taken 80% or more of the prescribed number of tablets, and poor if less than 80% tablets were taken during treatment.

### Outcome measures

Number of patients cured at 8 weeks with the two regimens was the primary outcome measure. Secondary outcome measures included number of patients cured at 4 weeks and frequency of adverse events. In addition, severity of itching was noted on a 0 (no itching) to 10 (most severe itching) visual analogue scale at baseline and subsequent follow-ups.

### Statistical analysis

For the baseline variables, mean and standard deviation (SD) or median and interquartile range (IQR, IQ1- IQ3) were calculated depending on the distribution of data. Cure rates and compliance were compared by Fisher exact test. P values less than 0.05 were considered significant. All P vales are two-tailed. When appropriate, 95% confidence intervals (CI) were calculated. Intention to treat analysis was performed for the effectiveness data. Compliance data were analysed on per protocol basis. Denominator for calculating compliance was the total number of patients who were cured and those who came for follow ups up to 8 weeks but were not cured.

## Results

### Baseline characterises of the patients

Patients in the 2 groups were comparable at baseline with regard to age, gender, weight, duration of tinea, and body surface area affected (Table 1). Seventy nine had tinea corporis et cruris, 31 tinea corporis et cruris et faciei, 6 tinea corporis, 9 tinea cruris, 4 tinea faciei, and 3 had tinea cruris et faciei. Ten patients had received oral antifungal treatment in past (none received terbinafine or griseofulvin in past 1 month). Fifty patients in each group had applied some topical preparation in the past (47 had used topical steroid in any form in T and 46 in T+G group).

**Table 1.** Baseline characteristics of the patients.

|  | Terbinafine<br>n=66 | Terbinafine+griseofulvin<br>n=66 |
| --- | --- | --- |
| Age (years)<br>(mean±SD) | 26.5 ± 11.2 | 23.6± 9.1 |
| Female patients, n (%) | 17 (25.8) | 21 (31.8) |
| Duration (months),<br>median (IQR) | 4 (2.5-12) | 4 (3-10.8) |
| Weight (kilograms)<br>(mean±SD) | 64.0± 11.7 | 60.2± 12.5 |
| Body surface area (BSA)<br>affected | 3.8± 1.7 (mean±SD) | 3 (2-6) (median, IQR) |
| Comorbidities | Vitiligo 1<br>Hypothyroidism 2 | Vitiligo 1 |

Normally distributed data are presented as mean±SD. Data which are not normally distributed are presented as median and IQR. IQR, interquartile range; SD, standard deviation.

### Cure rates at 4 weeks

At 4 weeks, none of the patients were cured in any group.

### Cure rates at 6 weeks

At 6 weeks, 1(1.5%) and 4 (6.1%) patients were cured in T and T+G groups, respectively. The cure rates were not significantly different (P=0.417) (Fisher exact test) (Table 2).

**Table 2.** Number of patients cured at 4, 6 and 8 weeks with the two treatments.

| Treatment | 4 weeks | 6 weeks<sup>a</sup> |  | Up to 8 weeks<sup>b</sup> |  |
| --- | --- | --- | --- | --- | --- |
|  | Patients cured (%), ITT | Patients cured (%), ITT | 95 % CI (%) | Patients cured (%), ITT | 95 % CI (%) |
| Terbinafine | 0 | 1 (1.5) | <0.01 to 8.88 | 17 (25.8) | 16.67 to 37.51 |
| Terbinafine+griseofulvin | 0 | 4 (6.1) | 1.94 to 15.01 | 19 (28.8) | 19.21 to 40.70 |

<sup>a</sup>P=0.417, <sup>b</sup> P=0.845 (Fisher exact test). CI, confidence interval; ITT, intention to treat analysis.

### Cure rates at 8 weeks

At 8 weeks, 17 (25.8%) and 19 (28.8%) patients were cured in T and T+G groups, respectively. Cure rates at 8 weeks with the two treatments were not significantly different (P=0.845) (Fisher exact test) (Tables 2, 3).

**Table 3.** Comparison of cure rates at 8 weeks with the two treatments.

| Number of patients cured with terbinafine, ITT (n=66) (%) | Number of patients cured with terbinafine +griseofulvin, ITT (n=66) (%) | P<sup>a</sup> | Difference | 95% CI of difference (%) |
| --- | --- | --- | --- | --- |
| 17 (25.8) | 19 (28.8) | 0.845 | 3.03 % | -12.16 to 18.22 |

<sup>a</sup>Fisher exact test

### Number needed to treat

Number needed to treat (NNT) for the experimental group T+G was 33 (versus T) calculated based on cure rates at 8 weeks (Table 4).

**Table 4.** Number needed to treat and absolute risk reduction with terbinafine plus griseofulvin versus terbinafine.

| **Treatment** | **NNT** | **ARR** | **95% CI of ARR (%)** |
| --- | --- | --- | --- |
| Terbinafine+griseofulvin | 33 | 3.03% | -12.16 to 18.22 |

Absolute risk reduction of treatment failure versus terbinafine at 8 weeks.
ARR, absolute risk reduction; CI, confidence interval; NNT, number needed to treat.

### Severity of itching

Severity of itching decreased in all treatment groups as the treatment progressed and itching was absent in all cured patients at the time of cure (Table 5).

**Table 5.**
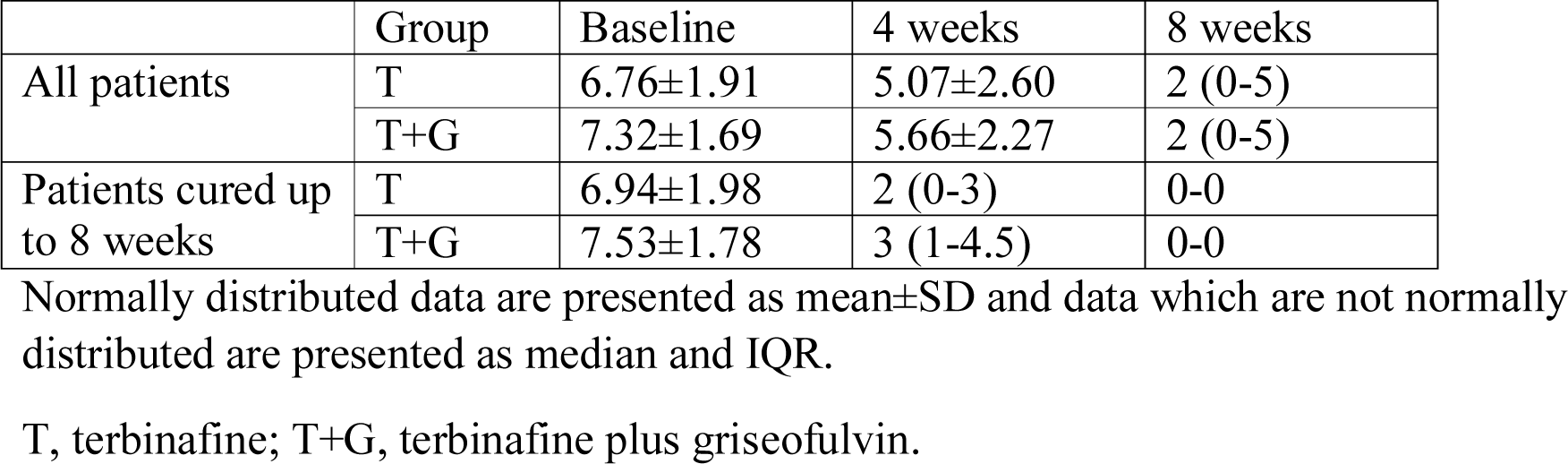
Severity of itching.

|  | Group | Baseline | 4 weeks | 8 weeks |
| --- | --- | --- | --- | --- |
| All patients | T | 6.76±1.91 | 5.07±2.60 | 2 (0-5) |
|  | T+G | 7.32±1.69 | 5.66±2.27 | 2 (0-5) |
| Patients cured up to 8 weeks | T | 6.94±1.98 | 2 (0-3) | 0-0 |
|  | T+G | 7.53±1.78 | 3 (1-4.5) | 0-0 |

Normally distributed data are presented as mean±SD and data which are not normally distributed are presented as median and IQR.
T, terbinafine; T+G, terbinafine plus griseofulvin.

### Compliance

Sixty two out of 63 patients and 59 of 65 patients had good compliance in T and T+G groups, respectively. The difference was not significant (P=0.115).

### Patients’ satisfaction with treatment outcome

All patients, who were cured, were fully satisfied with the outcome.

### Adverse events

No adverse events were detected on investigations and none were reported by the patients.

## Discussion

Present study was conducted in view of unprecedented changes in the epidemiology, clinical features and treatment responsiveness of tinea infections in recent past in India.^1,2^ Recent data show that terbinafine, once a highly effective drug, now has abysmal cure rate in tinea corporis, tinea cruris and tinea faciei.^3,4^ Furthermore, recent data show that effectiveness of other oral antifungal drugs (fluconazole, griseofulvin, itraconazole) has also declined in India.^4^ These results confirm the general impression among Indian dermatologists of decreased effectiveness of antifungal drugs in current epidemic of dermatophytosis in India.

Terbinafine has been considered to be the oral antifungal drug of choice for dermatophytosis worldwide in view of its fungicidal action, high cure rate, minimal drug interactions and a general lack of serious adverse effects. However, this is no longer the case in the current epidemic of altered dermatophytosis in India, which is characterized by altered morphology, extensiveness and chronicity and frequent relapses within weeks of apparent cure.^8^ It is obviously important that new treatment methods are tested for their effectiveness in tinea infections in the current situation.

In the present study, we tested the hypothesis that using a multidrug therapy regimen may produce better treatment outcome in tinea compared to single agent in India. In the past, combination of antifungal agents with complementary targets within the fungal cells has been used for invasive fungal infections.^9^ Terbinafine and griseofulvin were selected as the two antifungal agents because they have different mechanisms of action. Effectiveness of oral terbinafine daily (T) (active control) was compared with terbinafine plus griseofulvin daily (T+G) in tinea corporis, tinea cruris and tinea faciei within a randomized pragmatic open design.

At 4 weeks, none of the patients were cured in either group. Cure rates were 1.5% versus 6.1% at 6 weeks, and 25.8% versus 28.8% at 8 weeks, with terbinafine and terbinafine plus griseofulvin regimens, respectively. The cure rates at all time points were not significantly different between the groups, and were disappointingly low with both treatments. Considering the cure rates at 8 weeks, number needed to treat (NNT) for terbinafine plus griseofulvin regimen was 33, meaning thereby that 33 patients would have to be treated for one additional patient to be benefitted versus terbinafine alone, thus showing the futility of adding griseofulvin.

Compliance of the patients of the two groups was similar. No adverse events were detected on investigations and none were reported by the patients during the study.

A limitation of the present study is that it was not blinded.

To conclude, data presented herein show the futility of combining griseofulvin and terbinafine for treatment of patients with tinea corporis, tinea cruris and tinea faciei in the current epidemic of altered dermatophytosis in India. Urgent need of better treatment options cannot be overemphasized.

## Data Availability

No

---

**Figure 1.**
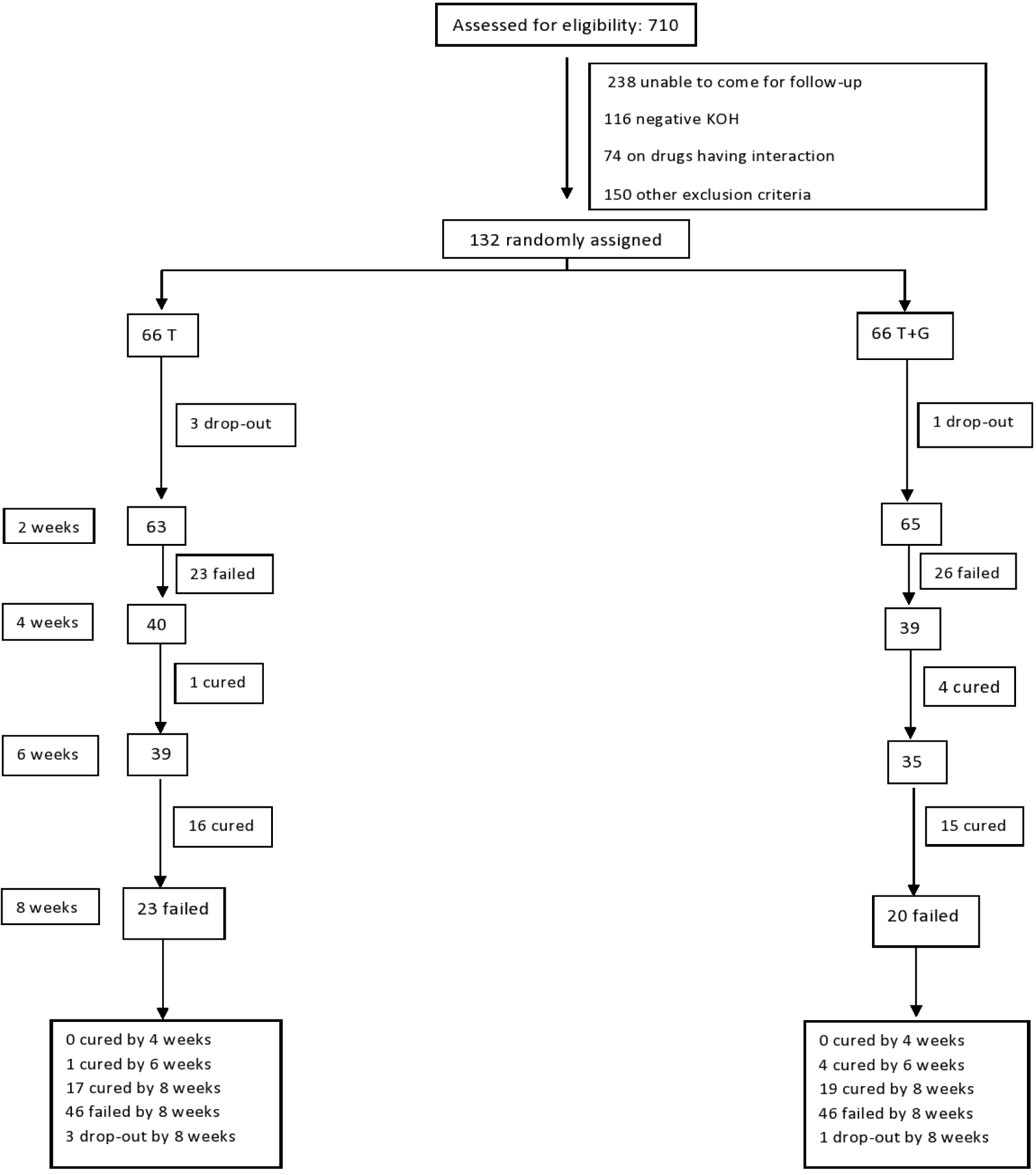
Flow diagram of the clinical trial. KOH, 20% potassium hydroxide microscopy; T, terbinafine; T+G, terbinafine plus griseofulvin.

## Notes

### Competing Interest Statement

The authors have declared no competing interest.

### Clinical Trial

CTRI/2019/06/019778

### Funding Statement

Banaras Hindu University
No other funding

### Author Declarations

All relevant ethical guidelines have been followed and any necessary IRB and/or ethics committee approvals have been obtained.

Any clinical trials involved have been registered with an ICMJE-approved registry such as ClinicalTrials.gov and the trial ID is included in the manuscript.

